# Physiotherapist-Led Stroke Rehabilitation in India: Preliminary Results from the Karnataka Brain Health Initiative (KaBHI) Model

**DOI:** 10.64898/2026.01.03.26343373

**Authors:** Mayank Sharma, Ganagarajan Inbaraj, Aparna Venugopal, Suvarna Alladi, Faheem Arshad, Girish B Kulkarni, Girish N Rao, Rajani Parthasarathy, Rehan Shahed, V Selva Ganapathy

## Abstract

**Background:** Post-stroke disability contributes substantially to long-term functional limitation, yet access to rehabilitation remains inequitable in low and middle-income countries (LMICs), where workforce and service delivery constraints are pronounced. Global priorities, including WHO’s Rehabilitation 2030 agenda, emphasize integrating scalable rehabilitation into health systems and decentralizing services beyond tertiary-centers. Evidence supports home and community-oriented approaches, including caregiver-mediated exercise, that can achieve gains comparable to centre-based therapy when dose is matched. The Karnataka Brain Health Initiative (KaBHI) is a public-sector model strengthening neurological care pathways through district Brain Health Clinics (BHCs).

**Objective:** To evaluate short-term changes in balance, mobility, and global disability among community-dwelling stroke survivors completing the KaBHI physiotherapy programme.

**Methods:** This prospective, single-group pre–post evaluation included adults aged 18–90 years enrolled at KaBHI BHCs (April–December 2024). Participants received a structured six-week balance and mobility programme prescribed by physiotherapists and implemented with caregiver support and follow-up within routine-care. Outcomes assessed at baseline (day 0) and post-intervention (day 45) were the Berg Balance Scale (BBS), Timed Up and Go (TUG), and Modified Rankin Scale (mRS).

**Results:** Among 199 participants (mean age 54.9 ± 14.5 years; 71% male), BBS improved from 28.9 to 37.2 (p < 0.001), TUG improved from 22.7 s to 19.8 s (p < 0.001), and mRS decreased from 3.30 to 2.45 (p < 0.001). Clinically meaningful improvement was common: 51% achieved ≥7-point gains on BBS, and 70% improved by ≥1 mRS grade. No programme-related serious adverse events were reported.

**Conclusions:** In a real-world public health setting, a physiotherapist-led, caregiver-supported KaBHI rehabilitation model delivered through district BHCs demonstrated significant short-term improvements in balance, mobility, and disability. These findings align with evidence supporting decentralized, home-/community-oriented, caregiver-mediated rehabilitation and support KaBHI as a potentially scalable strategy to strengthen stroke rehabilitation capacity in resource-constrained settings; controlled and longer-term evaluations are warranted.

## Introduction

Stroke is a leading cause of disability worldwide. Approximately 87% of stroke-related disability occurs in low- and middle-income countries (LMICs)^1^, yet rehabilitation services in these settings are grossly inadequate. On average, LMICs have only about 10 rehabilitation professionals per million people, compared to hundreds per million in high-income countries.^1,2^ A survey by the World Stroke Organization found that specialized stroke rehabilitation units were present in only 18% of responding LMICs versus 91% in high-income countries.^3^ This enormous service gap leaves the majority of stroke survivors in LMICs without access to needed therapy, contributing to poorer functional outcomes and higher long-term disability.^1^ In countries like India, stroke incidence is rising even as rehabilitative care remains concentrated in urban tertiary centers that are financially and geographically inaccessible for many patients. Rural and semi-urban communities face a lack of trained therapists and infrastructure, leading to unmet needs in post-stroke care.^4^

Recognizing rehabilitation as integral to quality healthcare, the World Health Organization’s “Rehabilitation 2030” call to action urges countries to integrate rehab services at all levels of the health system.^5^ Innovative, community-focused models are needed to extend stroke recovery support beyond hospital walls. Prior studies in LMICs have highlighted strategies such as home-based exercise programs, task-shifting to community health workers or family caregivers, and telerehabilitation as promising approaches to improve access.^1^ The Brazil’s nationwide Family Health Strategy (FHS) uses over 265,000 community health workers to provide basic health services to 67% of the population at the community level.^6^ This primary care network has been associated with significant improvements in health outcomes, including reduced stroke-related hospitalizations and mortality, by bringing follow-up care into patients’ homes. Similarly, in Bangladesh and other South Asian LMICs, where formal stroke rehab services are limited, community-based rehabilitation programs and caregiver training have emerged to fill the gap.^7^ These initiatives align with global recommendations that policymakers invest in expanding community rehabilitation and strengthening workforces to overcome barriers in stroke care.

Within India, the Karnataka Brain Health Initiative (KaBHI) represents a first-of-its-kind, state-led effort to integrate stroke rehabilitation into the public health system at scale. Launched in 2022 by the Government of Karnataka in collaboration with the National Institute of Mental Health and Neurosciences (NIMHANS), KaBHI established a network of Brain Health Clinics (BHCs) at district hospitals across the state.^8^ Each BHC is staffed by a multidisciplinary team (neurologist, physiotherapist, speech therapist, psychologist, nurse, and coordinator) to provide comprehensive care for neurological disorders, including stroke. KaBHI employs a hub-and-spoke model: the district BHC serves as the hub where stroke survivors receive initial assessment and individualized rehabilitation prescriptions from a physiotherapist, while the “spokes” extend into the community via home-based exercise programs supervised by caregivers. This model leverages existing health infrastructure and family engagement to overcome geographical and resource constraints, bringing stroke therapy into patients’ homes. Importantly, KaBHI’s physiotherapy program is delivered free-of-cost through the public system, reducing the financial burden on families and aiming to improve equity in rehab access.

The present study reports the preliminary outcomes of the KaBHI physiotherapist-led stroke rehabilitation program after six weeks of intervention. We aimed to evaluate real-world changes in balance, mobility, and functional independence among community-dwelling stroke survivors who participated in the home-based program. We hypothesized that even a short-term, low-cost intervention delivered via a task-shifting approach (physiotherapist guidance with caregiver implementation) could yield measurable functional improvements. Using standardized outcome measures, we assessed pre- to post-program changes and examined the proportion of participants achieving clinically meaningful improvements. Additionally, we describe the program design and implementation in detail, following STROBE and TIDieR guidelines, to inform replication in similar low-resource settings. By contextualizing our findings with national and international rehabilitation initiatives, we discuss the broader implications of the KaBHI model for strengthening neurorehabilitation services in LMIC health systems.

## Methods

### Study Design

We conducted a prospective single-group interventional study (pre–post design) to evaluate short-term functional outcomes of the KaBHI stroke rehabilitation program. All participants received the same standardized intervention as part of routine care at KaBHI BHCs. Outcome assessments were performed at the beginning of the program (baseline, Day 0) and after completing the 6-week intervention (follow-up, Day 45). This design corresponds to a practice-based implementation evaluation of a new service, and it is reported in accordance with STROBE guidelines for observational studies and the TIDieR checklist for intervention description.

### Setting

The Karnataka Brain Health Initiative was implemented state-wide in Karnataka, India, and by 2024 it had established 33 Brain Health Clinics embedded in district and medical college hospitals. These clinics act as regional hubs for neurological care under the state public health department, supported technically by NIMHANS. Each BHC has at least one trained physiotherapist responsible for stroke rehabilitation. After a stroke survivor is discharged from acute care or identified in the outpatient setting, they are referred to the BHC for follow-up. At the BHC, the physiotherapist assesses the patient and enrolls them into the home-based rehabilitation program if appropriate. This hub-and-spoke model ensures that while specialist input is available at the district hospital hub, the actual rehabilitation exercises are carried out at the patient’s home (the spokes) with periodic guidance. This design brings rehabilitation into rural and semi-urban communities that previously had little access to such services. All intervention and follow-up described took place between April and December 2024 across the network of BHCs.

#### Participants

Stroke survivors attending KaBHI Brain Health Clinics (BHCs) during the study period were screened for eligibility, and consecutive sampling was used to enroll all patients who met the study criteria. Eligible participants were adults aged 18–90 years with a physician-confirmed diagnosis of stroke (ischemic or hemorrhagic), who were medically stable, residing in the community, and able to undertake a home-based exercise programme either independently or with caregiver assistance. Inclusion further required completion of the structured six-week KaBHI physiotherapy programme, with both baseline (day 0) and follow-up (approximately day 45) outcome assessments documented. Participants were excluded if they had severe cognitive impairment or receptive aphasia that prevented understanding of instructions, unstable medical conditions that could make exercise unsafe (uncontrolled cardiac disease), or any clinical or safety factor that precluded participation in home exercise even with caregiver support, such as extreme frailty or a very high fall risk requiring constant professional supervision. Written informed consent was obtained from all participants prior to enrolment. Where needed, caregivers most commonly family members were identified and oriented to support home practice and safety monitoring. The enrolled cohort represented a broad range of stroke chronicity, including acute, subacute, and chronic phases, reflecting the BHC role in serving both recent post-discharge patients and longer-term stroke survivors seeking rehabilitation.

### Intervention: Home-Based Physiotherapy Program

The KaBHI physiotherapy program was designed to be a low-cost, scalable intervention targeting balance and mobility deficits common after stroke. It aligns with evidence that task-specific, repetitive training can improve functional outcomes, and addresses known barriers in LMICs by using minimal equipment and engaging family support.^1^ The program content was informed by standard stroke rehabilitation guidelines and adapted to the home environment by KaBHI physiotherapy specialists.

#### Intervention Components

Each participant underwent a six-week home exercise regimen focusing on: (1) Static and dynamic balance training, (2) Lower-limb strengthening, and (3) Functional mobility practice (Table 1). Each exercise session was intended to last 30 minutes, comprising a warm-up, targeted balance/strength drills, and a cool-down stretch.

**Table 1:**
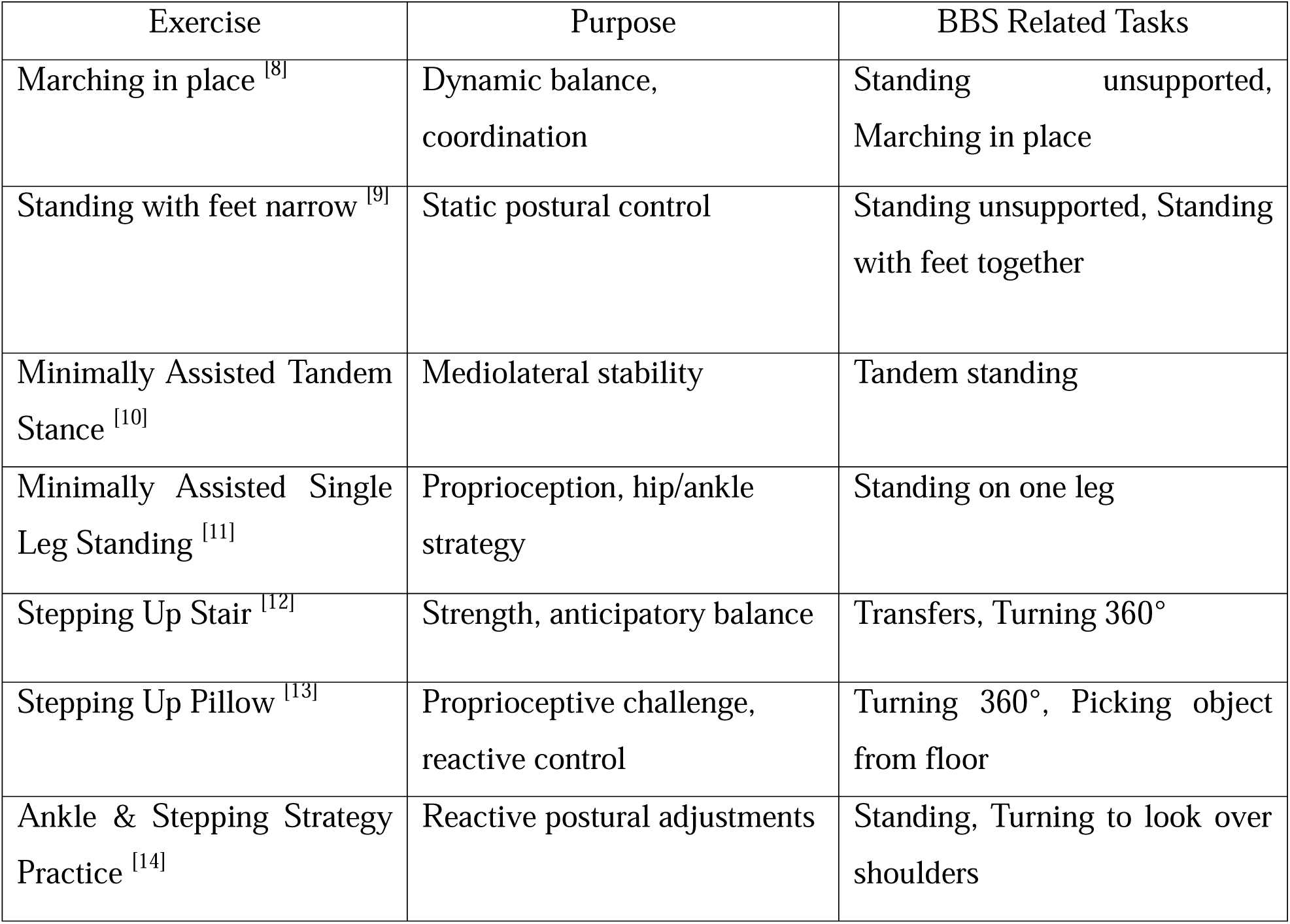
Key exercises used in the KaBHI balance training protocol.

| Exercise | Purpose | BBS Related Tasks |
| --- | --- | --- |
| Marching in place <sup>[8]</sup> | Dynamic balance, coordination | Standing unsupported, Marching in place |
| Standing with feet narrow <sup>[9]</sup> | Static postural control | Standing unsupported, Standing with feet together |
| Minimally Assisted Tandem Stance <sup>[10]</sup> | Mediolateral stability | Tandem standing |
| Minimally Assisted Single Leg Standing <sup>[11]</sup> | Proprioception, hip/ankle strategy | Standing on one leg |
| Stepping Up Stair <sup>[12]</sup> | Strength, anticipatory balance | Transfers, Turning 360° |
| Stepping Up Pillow <sup>[13]</sup> | Proprioceptive challenge, reactive control | Turning 360°, Picking object from floor |
| Ankle & Stepping Strategy Practice <sup>[14]</sup> | Reactive postural adjustments | Standing, Turning to look over shoulders |

#### Delivery and Mode

The programme was initiated with an in-person session at the district Brain Health Clinic (BHC), during which the physiotherapist assessed the participant, demonstrated the prescribed exercises to both the participant and caregiver, provided an illustrated instruction booklet in the local language, and confirmed safe performance of each activity. During this session, caregivers were trained as co-therapists to supervise home practice, provide physical support when needed, and maintain an exercise log. Thereafter, participants were instructed to perform the exercises at home on most days of the week (at least five days/week) for six weeks under caregiver supervision. In addition to home practice, participants were scheduled to attend the BHC once weekly for six consecutive weeks for supervised review, progression of exercise difficulty, reinforcement of technique, and safety monitoring. Between visits, participants could contact the physiotherapist for concerns, and telephone follow-up was used as needed to address adherence barriers or emerging symptoms. At each weekly review, the physiotherapist assessed progress, reviewed the exercise log, asked about difficulties or adverse events, and modified the programme when indicated. Overall, this delivery format represents a pragmatic hybrid model that combines structured, facility-based physiotherapist oversight with caregiver-supported home training, optimising adherence and safety while remaining feasible within a district public health service.

#### Materials and Resources

The intervention was intentionally low-tech. Exercises used common household items for support (a chair for seated/standing support, a step or low stool for step training, a cushion to simulate unstable surfaces). No specialized equipment was needed. Each participant received a printed handout with simple diagrams and instructions for each exercise, and a diary to check off daily sessions. The caregiver’s role was to ensure safety, help count repetitions, and note any concerns. This caregiver-supported approach is a form of task-shifting that leverages informal care resources – an important strategy in LMIC contexts where healthcare workforce is limited.^1^

#### Intervention Fidelity

All physiotherapists delivering the program were trained in KaBHI workshops to standardize the exercise protocol and follow-up procedures. They used a checklist during initial training to cover all exercises and safety points. While direct observation of home sessions was not feasible, the combination of caregiver logs and follow-up calls provided some indication of fidelity. Given this was a real-world implementation, minor deviations in how exercises were done at home were expected; the focus was on ensuring at least a core set of balance exercises were practiced regularly by each patient.

### Outcomes and Assessments

Primary outcomes were change in balance, mobility, and global disability, as measured by three widely used clinical scales. Assessments were conducted at baseline (Day 0) before starting the program, and at post-intervention (Day 45), typically during a follow-up visit to the BHC or occasionally at home for those unable to travel.

#### Berg Balance Scale (BBS)

A 14-item performance test evaluating static and dynamic balance (score range 0–56, higher scores = better balance). Each participant was guided through tasks like standing up, transfers, reaching, turning, and single-leg standing; each task is scored 0–4. The minimal clinically important difference (MCID) on BBS is generally considered ∼7 points in stroke populations for noticeable functional improvement.

#### Timed Up and Go (TUG) Test

A timed mobility test where the patient stands up from a chair, walks 3 meters, turns, and sits down. The time in seconds is recorded; shorter times indicate better functional mobility. A TUG <10 s is normal for healthy adults, while >30 s indicates high fall risk in frail elders. Many stroke survivors are in between these extremes; changes of ∼2–3 seconds can signify meaningful mobility improvement.

#### Modified Rankin Scale (mRS)

A clinician-rated scale of global disability and independence (scores range from 0 = no symptoms, to 5 = severe disability bedridden; 6 denotes death, which was not applicable here). We focused on mRS 0–5 among survivors. A decrease by 1 grade (i.e., from 4 to 3) is often considered a clinically significant gain in independence. mRS was determined by interview, considering the patient’s ability to perform daily activities with or without assistance.

All outcome assessments were performed by the BHC physiotherapists who were providing care, using standardized protocols to administer each test. Where possible, the same therapist assessed a given patient at baseline and follow-up to maintain consistency. To minimize bias, therapists were instructed to record objective measurements (time in seconds, task completion ability) without providing excessive encouragement that could differ between baseline and follow-up.

Additionally, basic demographic and clinical data were recorded: age, sex, stroke type (infarct or hemorrhage, when known), stroke chronicity (time since stroke and categorized phase: acute <1 month, subacute 1–6 months, chronic >6 months), use of assistive device (cane/walker), and common comorbidities (hypertension, diabetes). These characteristics were noted to contextualize the sample and explore any baseline factors that might influence outcomes.

### Data Collection and Analysis

All data were recorded in structured clinical forms at each BHC and then entered into a centralized database. For analysis, we used IBM SPSS Statistics (v25). Continuous variables were summarized as mean and standard deviation (SD) if approximately normally distributed, or median and interquartile range (IQR) for skewed distributions. Categorical variables were summarized as counts and percentages. Normality of outcome score distributions was tested with the Shapiro–Wilk test, which suggested non-normal distributions for some measures (given ordinal or skewed nature of scores). Therefore, non-parametric tests were chosen for the main pre–post comparisons. We used the Wilcoxon signed-rank test (a non-parametric analog of the paired t-test) to evaluate the change from baseline to 6 weeks for BBS, TUG, and mRS. A two-tailed p-value <0.05 was considered statistically significant for these analyses. In addition to p-values, we calculated effect sizes for the Wilcoxon test (denoted by the rank biserial correlation *r*) to quantify the magnitude of change. No imputation was done for missing data; only participants with both baseline and follow-up measurements on a given outcome were included in the analysis.

### Ethical Considerations

The study was approved by the Institutional Ethics Committee of NIMHANS (No. NIMHANS/36th IEC [BS & NS DIV. 2022]). As this was part of a government health program evaluation, all procedures were conducted in line with the Declaration of Helsinki and local ethical guidelines. Each participant (or an appropriate surrogate for those with communication difficulties) provided informed consent after explanation of the program and data collection. Participation in the rehabilitation program was voluntary and patients could withdraw at any time without affecting their routine care. Because the intervention was of minimal risk (exercise therapy), precautions were taken to ensure safety: participants and caregivers were instructed to perform exercises on a flat surface, use a wall or chair for support when needed, and to skip any exercise that caused pain or dizziness. They were given a direct phone contact to the BHC physiotherapist to report any adverse events or concerns. No serious adverse events occurred during the program. To protect confidentiality, only de-identified data were used in analyses, and individual identities are not revealed in this report.

## Results

### Participant Characteristics

A total of 199 stroke survivors met the inclusion criteria and completed the 6-week follow-up assessment. Baseline characteristics are summarized in Table 2. The cohort’s mean age was 54.9 ± 14.5 years (range 18–85), and 142 (71%) were male. Just over half (55%) resided in rural districts, reflecting KaBHI’s outreach beyond urban centers. Time since stroke at enrolment varied widely (median 5 months, IQR 2–12). Participants were almost evenly distributed across stroke chronicity phases: 23% acute (<1 month post-stroke), 39% subacute (1–6 months), and 39% chronic (>6 months post-stroke). Subgroup trends suggested similar improvement magnitudes across acute, subacute, and chronic groups, though slightly larger absolute gains were observed in acute cases.Most patients had sustained ischemic strokes (exact stroke type was not systematically recorded but the majority were presumed ischemic based on clinical records). At baseline, functional status was generally low: the mean BBS was 28.9 (SD 16.1), indicating moderate balance impairment (for context, a score <45/56 often signifies significant fall risk). The mean TUG time was 22.7 ± 8.3 seconds; notably, 18% of tested individuals took >30 seconds, falling into a high fall-risk category. The average mRS score was 3.3 ± 1.2, with a median of 3 (IQR 2–4), corresponding to moderate disability (requiring some help but able to walk unassisted for mRS 3, or more substantial assistance for mRS 4 in some cases). About 17.6% of patients used a cane or walker for ambulation at baseline. Common vascular risk factors were prevalent: 138 (69%) had hypertension and 60 (30%) had diabetes. These baseline data illustrate that the program engaged survivors with significant residual disability, many of whom were months post-stroke and living in rural communities where continued rehab options were previously scarce.

**Table 2:** Baseline characteristics of the stroke survivor cohort.

| Characteristic | Stroke survivor (n=199) |
| --- | --- |
| Age, years | 54.9 ± 14.5 (range 18–85) |
| Sex, n (%) | Male 142 (71.4%), Female 57 (28.6%) |
| Stroke type | Not systematically recorded (majority presumed ischemic) |
| Stroke phase at enrolment | Acute <1 month: 45 (22.6%) Subacute 1–6 months: 77 (38.7%) Chronic >6 months: 77 (38.7%) |
| Time since stroke, median | 5 months (IQR 2–12 months)<sup>#</sup> |
| Assistive device use | 35 (17.6%) using cane or walker at baseline |
| Berg Balance Scale (BBS) | 28.9 ± 16.1 (median 30,IQR 18–44) |
| Timed Up & Go (TUG), seconds | 22.7 ± 8.3 (median 20.1,IQR 17–28) |
| Modified Rankin Scale (mRS) | 3.3 ± 1.2 (median 3,IQR 2–4) |
| Comorbid hypertension | 138 (69.3%) |
| Comorbid diabetes mellitus | 60 (30.2%) |
| Residence | Rural: 109 (54.8%),Urban: 90 (45.2%) |

*Data are mean ± SD or n (%) unless otherwise indicated.*
*<sup>#</sup>Time since stroke was approximated from clinical records, exact onset dates unavailable for some participants.*

### Outcomes After 6 Weeks of Rehabilitation

#### Balance (BBS)

There was a significant improvement in balance ability after the intervention. Mean BBS score increased from 28.9 at baseline to 37.2 at 6 weeks (Δ= +8.3 points, *p* < 0.001 by Wilcoxon test). This large change is well above the conventional 7-point threshold for clinical significance in chronic stroke rehabilitation. In fact, 51% of participants improved their BBS by at least 7 points (and 32% improved by ≥10 points). At baseline only 14% of the cohort had BBS ≥45 (often considered the cut-off for low fall risk), whereas after the program 32% reached that level, effectively doubling the proportion of patients with near-normal balance. The effect size for BBS change was *r* = 0.83, indicating a robust magnitude of improvement.

#### Mobility (TUG)

Functional mobility and agility also improved. The average TUG time reduced from 22.7 seconds to 19.8 seconds post-intervention (mean change –2.9 s, *p* < 0.001). Median TUG improved from 20.1 to 18.0 seconds approaching stroke-specific minimal detectable change (MCID) values, which are typically reported in the range of 2.9–3.6 seconds. While a ∼3-second improvement may seem modest, it is noteworthy for a diverse cohort and was statistically significant with *r* = 0.82. Importantly, among those who were very slow at baseline (i.e., the 18% with TUG >30 s), most showed some decrease in time; a few individuals moved from above 30 s into the 20–30 s range, suggesting reduced fall risk. Faster walkers also improved speed, reflecting better confidence and strength. TUG has some measurement variability, but the consistent direction of change across the sample indicates a genuine mobility benefit from the exercises (which included sit-to-stand practice, stepping, and walking balance tasks).

#### Disability (mRS)

Global disability scores shifted towards greater independence. The mean mRS decreased from 3.30 to 2.45 (*p* < 0.001). In practical terms, this means the average patient moved from a status of “moderate disability; requires some help” to between “slight to moderate disability.” Notably, 70% of patients improved by at least 1 mRS grade, and 27% improved by 2 grades. The proportion of participants who were mildly disabled (mRS ≤2, able to handle activities of daily living independently) nearly doubled from 26% at baseline to 49% after the program. These shifts highlight that even within six weeks, meaningful functional gains were achieved for many. However, no patients achieved mRS 0 or 1 if they were ≥2 at baseline, as expected given residual deficits; and those with very high baseline disability (mRS 5) were few in number and remained dependent (none improved beyond mRS 4). No participant had mRS 5 at follow-up (the worst was 4), despite a few being 5 at baseline, reflecting no one remained completely bedbound after intervention (though some still required assistance). The proportion with high fall risk on TUG (>30 s) decreased from 18% to 11%. These clinically meaningful shifts reinforce the quantitative improvements observed. The effect size for mRS improvement was *r* = 0.78, also a large effect by conventional standards for an intervention (Table 3).

**Table 3:**
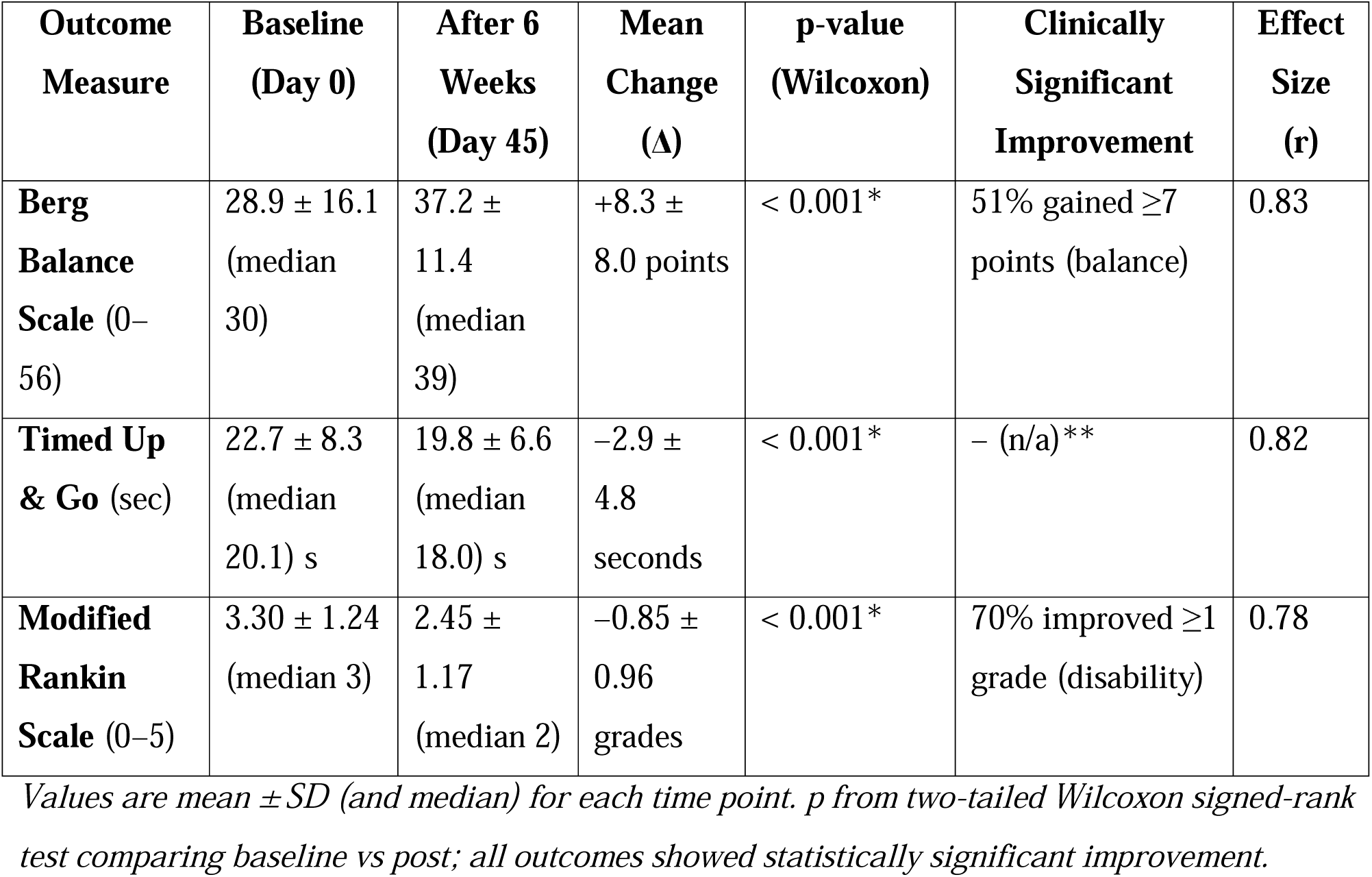
Pre- and Post-Intervention Outcomes for Balance, Mobility, and Disability.

| **Outcome Measure** | **Baseline (Day 0)** | **After 6 Weeks (Day 45)** | **Mean Change ($\Delta$)** | **p-value (Wilcoxon)** | **Clinically Significant Improvement** | **Effect Size (r)** |
| --- | --- | --- | --- | --- | --- | --- |
| **Berg Balance Scale (0–56)** | 28.9 ± 16.1<br>(median 30) | 37.2 ± 11.4<br>(median 39) | +8.3 ± 8.0 points | < 0.001* | 51% gained ≥7 points (balance) | 0.83 |
| **Timed Up & Go (sec)** | 22.7 ± 8.3<br>(median 20.1) s | 19.8 ± 6.6<br>(median 18.0) s | –2.9 ± 4.8 seconds | < 0.001* | – (n/a)** | 0.82 |
| **Modified Rankin Scale (0–5)** | 3.30 ± 1.24<br>(median 3) | 2.45 ± 1.17<br>(median 2) | –0.85 ± 0.96 grades | < 0.001* | 70% improved ≥1 grade (disability) | 0.78 |

*Values are mean ± SD (and median) for each time point. p from two-tailed Wilcoxon signed-rank test comparing baseline vs post; all outcomes showed statistically significant improvement.*

#### Adherence

Program adherence was high overall. Based on caregiver-maintained logs and patient self-report at follow-up, the median frequency of home exercise practice was 5 days per week (approximately 30 minutes per day). Eighty-five percent of participants practiced at least 4 days per week, and about 60% practiced 5–7 days per week consistently. Only a small minority (under 10%) fell below 3 days per week on average, usually due to intercurrent illness or travel that interrupted their routine. This high adherence can be attributed to the active follow-up and the integration of exercises into daily life with caregiver support. Participants often reported that having a family member involved kept them motivated and accountable. Adherence did not significantly differ by age or sex, though there was a slight tendency for those in the acute phase (recent stroke) to be more diligent (perhaps due to higher motivation for recovery), whereas some in the chronic phase had occasional complacency. Nonetheless, even chronic-phase survivors engaged well when given structure and support.

#### Safety

No program-related serious adverse events were reported during the 6-week period. In particular, no falls or injuries occurred while doing the prescribed exercises under caregiver supervision, according to participant reports. The proactive safety measures (starting exercises in therapist presence, caregiver standby, advice to skip difficult tasks) likely contributed to this excellent safety profile. This suggests that home-based rehabilitation, when properly guided, can be delivered with minimal risk even in remote settings.

### Adherence and Program Feasibility

As noted, adherence was good with a median of 5 exercise sessions per week. Qualitatively, patients and caregivers reported finding the exercises simple and feasible to do at home. Several patients noted improvements in confidence while walking (“I can walk to my field without fear now,” as one farmer reported) and caregivers observed better endurance in daily tasks.

No significant logistical problems were encountered in the six-week cycles besides minor challenges such as tracking some patients for follow-up if they missed appointments (which community health workers helped resolve in a few cases by visiting homes). All these indicators suggest the KaBHI model is feasible and acceptable to patients and providers in this resource-constrained setting.

## Discussion

This preliminary evaluation of the KaBHI physiotherapist-led stroke rehabilitation program demonstrates that structured home-based therapy with periodic professional guidance can yield significant functional gains among stroke survivors in a real-world public health setting. Over a 6-week period, participants showed marked improvements in balance, mobility, and overall disability status, with many achieving changes that are not only statistically significant but clinically meaningful for daily life. To our knowledge, KaBHI is the first state-funded program in an LMIC to combine district-level multidisciplinary clinics, caregiver-assisted home exercises, and tele-follow-up for stroke rehabilitation at this scale. The positive outcomes observed provide encouraging evidence that bridging the gap between hospital and home through a hub-and-spoke model can improve recovery trajectories for stroke survivors who would otherwise have limited rehabilitation access.

Our results align with findings from other studies that have tested home-based and community-oriented stroke rehab interventions.^9,10,11^ Prior research has indicated that task-specific exercise programs can improve balance and mobility even months or years post-stroke.^12^ What distinguishes KaBHI is the integration into the public system and reliance on task-shifting and caregiver engagement. In the global context, stroke rehabilitation in LMICs has been hindered by shortages of therapists, high costs, and geographic barriers.^1^ The KaBHI model addresses several of these determinants: it leverages government infrastructure to eliminate user fees, uses existing district hospitals to decentralize services, and enlists family members to extend the reach of care into the home. These elements align closely with recommendations from recent literature on improving post-stroke rehab access in LMICs. A 2025 scoping review identified home-based rehabilitation, caregiver-mediated interventions, and digital/telehealth support as three promising strategies to bridge the post-stroke care gap in resource-limited settings.^13^

KaBHI exemplifies how rehabilitation can be embedded into primary or secondary healthcare levels, which is a key goal of WHO’s Rehabilitation 2030 initiative.^5^ By operating through district hospitals and utilizing non-specialist caregivers, the program demonstrates feasibility at scale. Already 33 clinics are running under KaBHI, indicating reach across an entire state. This wide implementation hints at strong “reach” and “adoption” – two components of the RE-AIM framework (Reach, Effectiveness, Adoption, Implementation, Maintenance) often used to evaluate public health interventions. In terms of implementation, our results show that minimal resources can be leveraged effectively: aside from personnel time, the program required little equipment or funding, which bodes well for sustainability. Importantly, caregiver involvement effectively multiplies the workforce; family members become an extension of the rehab team. This task-shifting strategy has been successfully used in other health domains (HIV care, maternal health) and is now being recognized as critical for rehabilitation in LMICs.^14^ Our experience adds practical evidence that training and supporting informal caregivers can lead to safe and productive rehabilitation sessions at home. Policymakers in similar contexts can draw from KaBHI when aiming to expand stroke rehab services – leveraging community health worker networks or family engagement could amplify the impact of a limited number of professionals.

When comparing KaBHI to Brazil’s Family Health Strategy, one finds both parallels and differences. The FHS has a broader primary care mandate and has improved access to preventative and follow-up care nationwide, contributing to lower stroke hospitalization rates through risk factor management and early intervention.^15^ However, specific rehabilitation services within FHS remain inconsistently available; one study noted that only about 25% of Brazilian stroke survivors received any rehab, despite the universal health system.^16^ KaBHI, being a specialized program, ensures that stroke patients are actively provided rehab training and follow-up, rather than leaving it to chance. In essence, KaBHI adds a focused rehabilitation component that could complement primary care approaches. It highlights that beyond general healthcare access, dedicated rehab programs are necessary to address the long-term disabilities after stroke – a lesson applicable to many LMICs.

In South Asia, community-based rehabilitation (CBR) models have been promoted (often by NGOs) to serve persons with disabilities in their locales.^17^ Bangladesh, has CBR efforts where health workers or therapy aides visit communities to guide basic rehab, and family members are taught how to assist stroke survivors.^7^ These programs have reported improved patient satisfaction and perception of care, despite the paucity of formal therapists.^18^ KaBHI shares the ethos of CBR by making the community the center of care. The difference is that KaBHI is government-led and integrated into the health system, which could provide a more sustainable and standardized solution than purely NGO-run services. Our positive findings contribute evidence that government-run CBR-like initiatives can achieve measurable health outcomes. This supports calls for policymakers to invest in such models – as highlighted in a recent review urging expansion of community rehab and training as part of health policy in LMICs.^7^

The KaBHI model dovetails with the vision of WHO’s Intersectoral Global Action Plan for Epilepsy and Other Neurological Disorders 2022–2031, which advocates strengthening rehabilitation as part of integrated neurological care across all levels. It also answers the “call for action” to improve rehabilitation resources in health systems worldwide.^5^ By demonstrating an approach that can be embedded in existing public health infrastructure, our experience can inform WHO Rehab 2030 case studies and guidance. Essentially, KaBHI provides a working example of how to operationalize Rehabilitation 2030 principles in an LMIC context – through government leadership, task-sharing, capacity building of general health staff (training district physios in standardized care), and community empowerment via caregiver training.

Beyond the observed outcomes, this study demonstrates the feasibility of embedding an evidence-based, physiotherapist-led rehabilitation intervention into India’s public health system. Using the RE-AIM framework and Proctor et al.’s taxonomy of implementation outcomes, elements of reach (32 district hospitals), adoption (local physiotherapists and caregiver participation), and implementation (delivery using minimal resources) were evident.

To our knowledge, KaBHI is the first state-funded programme in a low- and middle-income country to combine district-level multidisciplinary clinics, caregiver-delivered home balance training, and low-bandwidth tele-support at scale. The intervention’s low equipment requirements, task-shifting approach, and integration within the National Programme for NCDs make it a potentially scalable template for sustainable stroke rehabilitation in resource-constrained settings.

The KaBHI initiative is an ongoing state-wide programme, and subsequent research cycles are already planned through the network of district Brain Health Clinics (BHCs). These upcoming evaluations will extend the current work across participating districts to compare KaBHI rehabilitation outcomes with usual care.

### Limitations

Despite its strengths, this study has several limitations. First, as an uncontrolled single-cohort study, we cannot definitively rule out that some improvements observed were due to spontaneous neurological recovery or placebo effects of attention. We mitigated this by focusing on short-term outcomes (6 weeks) where spontaneous improvements in chronic cases would be minimal, and by using objective measures. Still, without a comparison group, causation is inferred rather than proven. A related point is potential selection bias those who completed the program may be inherently more motivated or had family support, which could influence outcomes.

Some proportion of the improvement, particularly among acute/subacute survivors, may reflect natural recovery, although similar gains were also observed in chronic stroke.

Second, outcome assessments were performed by the treating physiotherapists, not blinded evaluators. This introduces risk of observer bias; therapists, optimistic about the program, might unconsciously encourage better performance at follow-up. We attempted to minimize this by using standardized instructions and by the objective nature of some measures (time in TUG). Still, the possibility of measurement bias means results should be interpreted with caution. Future studies ideally will have blinded assessments or use more objective endpoints (like sensor-based balance measures).

Third, the study duration was short. We report immediate post-intervention results, but do not know if these gains were maintained or translated into long-term reduction in falls or improved quality of life. Stroke recovery is an ongoing process; a 6-week snapshot misses later outcomes. We also did not formally measure quality of life or participation outcomes, which are important in chronic conditions. Similarly, cognitive outcomes or mood changes were not measured and could be relevant in a holistic evaluation.

Another limitation is that we did not systematically document fidelity of exercise performance or the exact content of each home session beyond self-reports. Thus, while adherence appears high, we rely on indirect reports. Use of exercise logs kept by caregivers could have reporting bias (overestimation of adherence). In the future, technology like wearable activity monitors or periodic video-supervision could give more objective adherence and fidelity data.

Our sample, though large for a single-center, is drawn from multiple districts with potentially heterogeneous implementation quality; we did not compare results across centers. There might have been variability depending on the physiotherapist’s experience or patient demographics (for example, maybe one district had mostly subacute patients, another mostly chronic). A stratified analysis could explore if acute vs chronic or rural vs urban had different outcomes, but our sample size for subgroups may limit power. We primarily report aggregate results here.

Finally, we have not conducted a formal cost analysis. While the program uses minimal additional resources, understanding the cost per patient and cost-effectiveness would be important for policy scale-up decisions. We also did not collect data on caregiver burden or satisfaction systematically – an important aspect since the model shifts work to families. Anecdotally, caregivers were positive about being involved, but future research should measure caregiver strain or empowerment outcomes.

## Limitations Summary

This study has several limitations. The absence of a control group limits causal attribution, as observed improvements cannot be definitively ascribed to the intervention alone. Outcome assessments were conducted by treating physiotherapists without blinding, introducing the possibility of assessor bias. Follow-up was short, so the durability of gains and longer-term outcomes such as falls and quality of life were not evaluated. Adherence was primarily self- or caregiver-reported, which may overestimate true exercise dose and fidelity in the absence of objective monitoring. Generalisability is also constrained because individuals with very severe deficits or major communication impairments were excluded, and the model assumes the availability of at least one willing caregiver, meaning patients without family support may not benefit similarly. Finally, we did not conduct a formal cost or process evaluation, including quantifying implementation costs, fidelity, or acceptability using standard metrics, although informal feedback suggested favourable acceptability. Despite these constraints, the findings provide useful real-world evidence from a low-resource public health setting and highlight priority areas to strengthen in subsequent programme phases.

## Future Directions

The encouraging findings from this preliminary evaluation justify more rigorous and broader research on the KaBHI rehabilitation pathway. Future work should prioritise controlled effectiveness designs, such as district-level quasi-experiments or stepped-wedge roll-outs, to quantify benefit over usual care while remaining feasible within a public programme. Longer follow-up (6–12 months) is needed to determine whether short-term gains are sustained and whether they translate into patient-important outcomes such as falls, readmissions, participation, return to work, and health-related quality of life. Subsequent studies should expand outcome assessment to include participation and patient-reported measures, and systematically capture caregiver outcomes, including burden, satisfaction, and perceived competence, given the model’s reliance on family support. Economic evaluation is also essential to inform scale-up, including programme delivery costs, downstream healthcare utilisation, and cost-effectiveness metrics. Technology-enhanced delivery should be tested pragmatically, comparing low-bandwidth follow-up with video-based telerehabilitation or simple mHealth tools, while accounting for digital access and literacy in rural populations. Because stroke recovery often requires multidisciplinary input, future KaBHI evaluations should examine integrated packages that combine physiotherapy with occupational therapy, speech and swallowing therapy, and psychological support, reflecting real-world BHC team capacity. Finally, the KaBHI stroke pathway can serve as a template for developing and evaluating similar district-integrated rehabilitation models for other neurological disorders managed within BHCs. Collectively, these steps will strengthen the evidence base, optimise implementation, and support policy decisions on embedding scalable rehabilitation pathways within national programmes such as NPCDCS.

## Conclusion

This pragmatic programme evaluation suggests that the KaBHI district-integrated, physiotherapist-led rehabilitation pathway is associated with clinically meaningful six-week improvements in balance, mobility, and disability among community-dwelling stroke survivors in a public-sector setting. By using Brain Health Clinics as hubs for assessment and progression and caregiver-supported home practice to extend exercise dose, the hub-and-spoke model offers a feasible approach to decentralizing rehabilitation where specialist services are limited. However, the uncontrolled design and short follow-up limit causal inference and do not address durability, falls, quality of life, or cost-effectiveness. Future controlled, longer-term evaluations with implementation and economic analyses are needed to confirm effectiveness across sites and to optimise delivery for people with severe disability or limited caregiver support.

## Data Availability

Availability of data and materials
Data supporting the findings of this study are available from the corresponding author on request.

## Declarations

### Ethics approval

This study was approved by the Institutional Ethics Committee at National Institute of Mental Health and Neurosciences, Ref: No. NIMHANS/36th IEC(BS&NS DIV)/2022 dated 08.07.2022. All procedures adhered to the Declaration of Helsinki and local ethical regulations. Written informed consent was obtained from all participants prior to enrolment. The exercise programme was designed to minimise risk, caregivers were involved in supervision, and participants were advised to modify or skip exercises that caused discomfort or instability.

### Clinical trial number

Not applicable, This study did not involve a clinical trial and therefore was not prospectively registered.

### Consent for publication

Not applicable

### Availability of data and materials

Data supporting the findings of this study are available from the corresponding author on request.

### Authors’ contributions

MS, GI, SA, and SG contributed to the study design development. MS, AV, contributed towards data collection, verification, and statistical analysis. MS, GI, SA, FA, GBK, GNR, RP, RS, and SG contributed to the manuscript preparation.

## Acknowledgements

We would like to thank the Scientific Advisory Group (SAG) of the Karnataka Brain Health Initiative (KaBHI) for their expertise extended towards the project: Suman P N Rao, Jeyaraj Durai Pandian, Giridhar R Babu, Subhash Kaul, Prashant Mathur, N. Sreekumaran Nair, Gururaj G.

